# Genetic and molecular analyses of candidate germline *BRIP1/FANCJ* variants implicated in breast and ovarian cancer

**DOI:** 10.1101/2023.07.03.23290133

**Authors:** Larissa Milano, Wejdan M. Alenezi, Caitlin T. Fierheller, Corinne Serruya, Timothée Revil, Kathleen K. Oros, Jeffrey Bruce, Dan Spiegelman, Trevor Pugh, Anne-Marie Mes- Masson, Diane Provencher, William D. Foulkes, Zaki El Haffaf, Guy Rouleau, Luigi Bouchard, Celia M.T. Greenwood, Jiannis Ragoussis, Patricia N. Tonin, Jean-Yves Masson

**Author notes:** Contributed equally to this study as co-first authors (Larissa Milano, Wejdan M. Alenezi). Corresponding authors. **CORRESPONDING AUTHORS**: Jean-Yves Masson, PhD, Faculty of Medicine, Université Laval, Québec, Canada, Senior scientist at the CHU of Quebec-Laval University Research Centre, Oncology axis, Quebec City, Québec G1R3S3, Canada, Patricia N. Tonin, PhD, Professor in the Departments of Medicine and Human Genetics, McGill University, Senior Scientist and Associate Leader, Cancer Research Program, Centre for Translational Biology The Research Institute of the McGill University Health Centre, E02.6217, 1001 Decarie Boulevard, Montreal, Quebec H4A 3J1.

## Abstract

Five rare variants in *BRIP1/FANCJ*, initially reported in ovarian (OC) or breast (BC) cancer cases by the adult hereditary cancer clinics, were investigated for their candidacy as clinically relevant variants. These variants were investigated genetically in a population exhibiting genetic drift and molecularly assayed for biological impact. Using in silico tools, population-based genetic databases and other resources, three of the five reported *BRIP1* variants were likely to be damaging: c.797C>T; p.Thr266Met, c.2087C>T; p.Pro696Leu and c.2990_2993delCAAA; p.Thr997ArgfsTer61. The carrier frequencies ranged from 0-0.7% in ancestry defined cancer groups comprised of 47 OC families, 49 hereditary breast and ovarian cancer syndrome families, 142 hereditary breast cancer syndrome families, 435 sporadic OC cases and 563 sporadic BC cases and 0-0.2% in 1025 population-matched controls. Multiple carriers of the same variants were identified in additional population-matched cancer cases. Of the five reported *BRIP1* variants, p.Thr266Met, p.Pro696Leu and p.Thr997ArgfsTer61, which were predicted to be damaging, conferred cellular sensitivity to mitomycin C and cisplatin unlike p.Ser139Ala and p.Ala406Ser. Collectively, our investigation implicates *BRIP1* c.797C>T; p.Thr266Met, c.2087C>T; p.Pro696Leu and p.Thr997ArgfsTer61 as deleterious variants in OC and BC.

## INTRODUCTION

*BRIP1* has been implicated as a hereditary breast cancer (BC) (1) and ovarian cancer (OC) predisposing gene (2). *BRIP1* was first reported as a BC predisposing gene in 2006 using a candidate gene approach involving hereditary BC (HBC) syndrome families that were not explained by germline pathogenic variants in *BRCA1* and *BRCA2* (1). Subsequent independent studies revealed no association of loss-of-function (LoF), pathogenic variants in *BRIP1* with BC though its role in BC risk remains equivocal (3–13). *BRIP1* was proposed as an OC predisposing gene in 2011 by a genome-wide association study of different cancer cases and controls, including OC (2). Subsequent studies consistently supported the association of pathogenic *BRIP1* variants with OC, suggesting that such variants in *BRIP1* play a role in conferring increased risk to OC (14). Carriers of pathogenic *BRIP1* variants in OC and BC cases are rare (15). Fewer than 1-5% of familial and sporadic OC or BC cases harbour pathogenic variants in *BRIP1,* which is significantly lower than the 20-80% of familial or 5-20% of sporadic OC or BC carriers of pathogenic *BRCA1* and *BRCA2* variants, depending on the population studied (3, 15, 16). While OC and BC cases harbouring pathogenic *BRIP1* variants are heterozygous (17), individuals homozygous or compound heterozygous for such variants are associated with the Fanconi anemia (FA) complementation group J (*FANCJ)*, a hereditary bone marrow failure syndrome exhibiting susceptibility to cancer (18–22).

*BRIP1*, also known as *BACH1* (BRCA1-Associated C-Terminal Helicase), was discovered in the context of elucidating the biological function of BRCA1 (23). BRIP1 and BRCA1 bind via their BRCA1 Carboxy-Terminus (BRCT) domains: one BRCT motif in BRIP1 and two in BRCA1 (23, 24). The BRCT domain in BRIP1 plays a critical role in its interaction with BRCA1 as a complex along with other proteins in mediating double-stranded DNA break repair by the FA and homologous recombination (HR) pathways (17, 18, 25, 26). Pathogenic variants in the BRCT domains of BRIP1 were shown to impact the repair of DNA double-strand breaks due to the loss of interaction between BRIP1 and BRCA1 (23, 27, 28). BRIP1 has also seven highly conserved DNA helicase motifs that it is essential for its catalytic activity in processing the repair of DNA inter– or intra-strand crosslinks (ICLs) via the FA-HR pathway (18, 23, 25, 26, 29, 30). Pathogenic variants in these helicase motifs have been shown to impact the repair of ICLs due to the loss of BRIP1 catalytic activity (27–29, 31, 32).

In this study, we investigated *BRIP1* variants for their candidacy as clinically relevant variants that were initially reported in BC and OC cases from adult hereditary cancer clinics in Canada. We applied a strategy involving the investigation of French Canadians (FC), a population known to exhibit unique genetic architecture due to genetic drift (3, 33, 34). The genetic analyses of this population has facilitated the characterization of pathogenic variants in known or candidate OC and BC predisposing genes (3). A small number of pathogenic variants in *BRCA1* and *BRCA2* (35, 36), one in *PALB2* (37), *RAD51C* (38) and *RAD51D* (38, 39) have been shown to have a higher allele frequency in FC OC and/or BC cases compared with population-matched controls. *BRIP1* was reported as a cancer predisposing gene based on the investigation of the germline variants in the Icelandic population, a well-documented founder population (40), where *BRIP1* c.2040_2041insTT was reported in 318 OC cases (2.36%) versus 0.41% of population-matched controls (2). *BRIP1*, however, has not yet been fully investigated in the FC population with only one early study reporting no clinically relevant variants in HBC and hereditary breast and OC (HBOC) syndrome families (41). We investigated candidate *BRIP1* missense variants using in silico tools selected for their best performance to predict their impact on gene function using a strategy recently applied to investigate missense variants in *RAD51C* and *RAD51D* identified in familial FC OC cancer cases (38, 39). We, then, investigated the carrier frequency of our candidates in genetically defined FC OC and BC cancer and control study groups.

We relate our findings to genetically characterized germline *BRIP1* variants identified in the Pan-Cancer OC and BC cases from The Cancer Genome Atlas (TCGA) (42) and cancer-free controls from the Genome Aggregation Database (gnomAD) (43). As our candidate *BRIP1* variants have not been characterized for their biological impact, we assayed in cell lines complemented with our BRIP1 variants and wild-type (WT) for cellular sensitivity to mitomycin C (MMC), cisplatin and poly (ADP-ribose) polymerase (PARP) inhibitors. Our genetic and molecular investigation of *BRIP1* candidate variants identified in a clinical context of the FC population facilitated the interpretation of candidate variants that are also relevant in other populations.

## MATERIALS AND METHODS

### Study groups

The study groups investigated in this report are described in **Table S1**. Information concerning *BRIP1* variants in OC or BC cases reported in clinical settings were obtained from adult hereditary cancer clinics in Canada. Study groups investigated for *BRIP1* variants were from participants selected from the following established biobanks: Banque de tissus et données of the Réseau de recherche sur le cancer of the Fond de recherche du Québec – Santé (RRCancer biobank) (rrcancer.ca); CARTaGENE (cartagene.qc.ca) (38, 44); Université de Sherbrooke-The Genetics of Glucose regulation in Gestation and Growth (Gen3G) (38, 45–47); McGill University-Montreal Neurological Institute (MNI) (38, 46, 48); and TCGA) (42). Clinical data (age of diagnosis, histopathology of cancer, disease stage, and tumour grade), genetic reports, and family history of cancer from selected cases were obtained from the respective biobanks and from adult hereditary cancer clinics. Information for each case was anonymized at source. For further protection of anonymity of participants in this study, we assigned a unique identifier (PT with four digits) to each case and further modified their respective pedigrees. Criteria for denoting FC ancestry is summarized in **Table S1**.

This project was conducted with approval and in accordance with the guidelines of The McGill University Health Centre Research Ethics Board (MP-37-2019-4783).

### Bioinformatic analyses of *BRIP1* variants reported for OC or BC cases of FC ancestry from adult hereditary cancer clinics

*BRIP1* variants initially reported in FC OC or BC cases were provided by adult hereditary cancer clinics (**Table S1**). These cases were also reported negative for pathogenic variants in *BRCA1* and *BRCA2*. These reported *BRIP1* variants were re-annotated using the canonical transcript NM_032043.3 (49). Variants retained for further analyses were those with a minor allele frequency (MAF) ≤0.01 in the general population in gnomAD v2.1.1. (43, 50) and being LoF) or missense variants classified as pathogenic, likely pathogenic or uncertain of significance (VUS) in ClinVar (51, 52) and/or by the American College of Medical Genetics and Genomics (ACMG) guidelines (53). Missense variants retained for further investigation were those predicted to be conserved and damaging at the RNA or protein level by at least one of the selected in silico tools as described previously (3, 38, 46, 47, 54–56). These in silico tools were selected based on their best performance (57–60). Three in silico tools for conservation prediction were applied: Genomic Evolutionary Rate Profiling (GERP++) v1.0 (61); Phylogenetic P value v4.2 of 100 vertebrates (PhyloP 100 way) (62) and PHAST Conservation v4.2 of 100 vertebrates (PhastCons 100 way) (63) with prediction scores of ≥2.0, ≥0.2 and ≥0.4, respectively. Five in silico tools predicting effect on splicing of transcript were applied: AdaBoost (ADA) v1.1 or Random Forest (RF) v1.1 with prediction scores of ≥0.4 (58), Maximum Entropy Estimates of Splice Junction (MaxEntScan) v2.0 (64), Human Splicing Finder (HSF) v3.1 (65) and Splice AI (66). Eight tools predicting effect on protein function were also applied: Rare Exome Variant Ensemble Learner (REVEL) v4.2 (67), Meta-analytic Logistic Regression (MetaLR) v4.2 (68), MetaRNN v4.2. (69), Variant Effect Scoring Test (VEST) v4.2 (70) with prediction scores of ≥0.4, Meta-analytic Support Vector Machine (MetaSVM) v4.2 (68), Eigen v4.2 (71) with prediction scores of ≥0.0, Combined Annotation Dependent Depletion (CADD) v1.6 (72) with prediction score of ≥20 and Protein Variation Effect Analyzer (PROVEAN) v4.2 (73) with prediction score of ≥-2.5.

### Determination of frequencies of carriers of *BRIP1* variants in defined FC cancer and control study groups

The carrier frequencies of our candidate *BRIP1* variants were investigated in FC study groups that have been extensively characterized in previous reports (**Table S1**). Briefly, *BRIP1* candidate variants were genotyped in peripheral blood lymphocytes (PBL) DNA from index cancer cases from five different FC groups: 47 OC, 49 HBOC and 142 HBC families (35, 36, 38, 46, 54, 74) as well as 435 sporadic OC and 563 sporadic BC cases (38, 46, 54, 75, 76), regardless of their status of pathogenic variants in *BRCA1* and *BRCA2* (**Table S1**). We genotyped samples using customized TaqMan^®^ (77), Sequenom iPLEX^®^ Gold (78) or Fluidigm^®^ SNP Type^™^ (79) genotyping assays (primers available upon request) as described previously (38, 46, 54, 56). Tumour DNA samples were genotyped where PBL DNA was not available. Carrier frequencies of *BRIP1* candidate variants were determined in population-matched controls by surveying 1025 sequencing-based data from: 433 from Gen3G, 422 from MNI and 170 from CARTaGENE; and surveying 8493 single nucleotide polymorphism (SNP) genotyping-based from CARTaGENE (38, 39, 45, 46, 54, 80, 81). For probes of variants not presented on the SNP arrays, pre-phasing and imputation were performed using Eagle2 with the Burrows-Wheeler transformation (82) through Sanger Imputation Services (sanger.ac.uk/tool/sanger-imputation-service/) using Haplotype Reference Consortium release (HRC.r1) v1.1 as a reference (83) as described previously (38, 46, 54). Pair-wise comparisons were performed of carrier frequencies of candidate variants in the different FC cancer groups versus sequencing-based controls. Two-tailed Fisher’s exact test was used to compare carrier frequencies in the cancer versus control groups where un-adjusted *P* values <0.05 for multiple testing was considered significant.

Additional carriers of candidate variants were identified in OC cases from two resources as described in **Table S1**: (1) whole exome sequencing (WES) data was available from 52 sporadic early-onset cancer cases diagnosed with high-grade serous ovarian carcinoma (HGSC) before the age of 50 years (38); and (2) targeted genotyping of PBL DNA or tumour DNA as described above from 534 recently recruited OC cases (38, 46, 84).

Candidate variants were verified in PBL DNA from the identified carriers by bidirectional Sanger sequencing using customized primers (primers available upon request) at the McGill Genome Center as previously described (38, 46, 54). Sequencing chromatograms were visually inspected for variant heterozygosity using 4Peaks v1.8. (nucleobytes.com/4peaks/) (The Netherlands Cancer Institute, Amsterdam, The Netherlands).

### Determination of carrier frequencies of candidate *BRIP1* variants identified in BC and OC cases and controls not selected for FC ancestry

We investigated *BRIP1* candidate variants in genetic data from OC and BC cases from the Pan-Cancer – TCGA (42) and non-cancer controls from the gnomAD v2.1.1. (43, 50), and both study groups were not selected for FC ancestry. Variant Call Format (VCF) files that were generated from WES data from the germline of 416 OC and 1072 BC Pan-Cancer – TCGA cases were downloaded as previously described (42, 54). Comma Separated Values (CSV) files that were generated from WES data from the germline of 134,187 cancer-free, non-Finnish European gnomAD v2.1.1. controls were directly downloaded from gnomad.broadinstitute.org. All variants in *BRIP1* were extracted from these files and annotated as previously described (3, 46, 54, 55). These variants were subjected to our filtering and prioritizing criteria as described previously (38). Variants with MAF >0.01 in the general population in gnomAD v2.1.1. were filtered out, and the remaining variants were prioritized for being: (1) LoF or missense variants predicted to affect splicing by at least 1 out of the four in silico tools as described above; (2) classified as pathogenic or likely pathogenic in ClinVar and/or by ACMG guidelines; (3) predicted to be conserved by at least one of the three selected in silico tools as described above; (4) predicted to be damaging at the level of the protein by at least six of the eight selected in silico tools as described above.

### Generation of constructs and cell lines for in cellulo assays of *BRIP1* variants

The pcDNA3-3xFlag-*BRIP1*-WT plasmid, expressing the Human BRIP1 Flag tagged with C-terminal 3X DDK tag, was kindly donated by Bob Brosh (NIA/NIH). The pcDNA3-3xFlag-*BRIP1* constructs harbouring one of our *BRIP1* variants were generated via site-directed mutagenesis using Q5^®^ Site-Directed Mutagenesis Kit (New England Biolabs, Canada) with primers listed in **Table S2**. The AAVS1 *BRIP1* WT or variant constructs were generated by amplification using the pcDNA3-3xFlag-*BRIP1* plasmids and primers listed in **Table S2**. Products were cloned into the AAVS1 vector in NotI/PspXI sites (85).

The U2OS (sarcoma derived cell line) and Hela (cervical carcinoma derived cell line) BRIP1 knock-out (KO) and control cells were kindly donated by Sharon Cantor (86, 87) and maintained in Dulbecco’s Modified Eagle Medium (DMEM) supplemented with 10% Fetal Bovine Serum (FBS) and 1% Penicillin-Streptomycin. BRIP1 KO cells were stably complemented using the AAVS1 genomic editing system (85). Briefly, cells were transfected with 4 μg of the AAVS1 construct containing either the WT or one of the BRIP1 variants, along with the 0.4 μg of the pZFN plasmid for 4h using Lipofectamine 2000 (Invitrogen, Canada). After 24h, transfected cells were selected with Gibco™ Geneticin™ Selective Antibiotic (G418 Sulfate) for 7 days. Established cell lines containing the BRIP1 variants were maintained in DMEM supplemented with 10% FBS, 1% Penicillin-Streptomycin and 0.5 mg/ml of G418 Sulfate.

### Drug sensitivity assays

The U2OS or Hela cells were seeded in triplicate assays into a Corning 3603 black-sided clear bottom 96-well microplate at a density of 2000 cells per well. MMC, cisplatin and PARP inhibitors (olaparib and talazoparib) sensitivity assays were, then, performed as previously described (88). Cells were treated with the indicated drugs for 4 days with concentrations ranging from 0 to 8 ng/ml for MMC, 0 to 60 μM for cisplatin, 0 to 2.5 μM for olaparib and 0 to 40 nM for talazoparib. The entirety of each well was imaged at 4x with Cytation 5 Cell Imaging Multi-Mode Reader and the Hoechst-stained nuclei were quantified using the Gen5 Data Analysis Software v3.03 (BioTek Instruments). Cell viability was expressed as percentage of survival of treated cells relative to vehicle-treated cells. Results represent the mean ± standard error of the mean (SEM) of at least 3 independent experiments, each performed in triplicate.

### Protein extraction and immunoblotting assays

Total soluble protein extracts and immunoblotting were performed as previously described (89). BRIP1 protein expression was detected using a polyclonal antibody (Sigma, #B1310). Anti-Tubulin (Abcam, #ab7291) served as the loading control. Anti-rabbit or anti-mouse IgG (Jackson ImmunoResearch) conjugated to horseradish peroxidase were used as secondary antibodies.

## RESULTS

### Candidate *BRIP1* variants were reported in FC probands with OC and BC by adult hereditary cancer clinics

We received information from three OC and four BC probands who tested positive for a *BRIP1* variant based on a 23-to-34 gene-panel testing for germline variants and copy number variants (CNV), except for two cases where CNV testing was done for only *BRCA1* and *BRCA2.* All seven probands self-reported FC ancestry. Medical genetic reports revealed that the four BC probands from independent families harboured a frameshift c.2990_2993delCAAA; p.Thr997ArgfsTer61 or missense variant c.415T>G; p.Ser139Ala in *BRIP1*, and the three OC probands each harboured a missense variant c.797C>T; p.Thr266Met, c.1216G>T; p.Ala406Ser or c.2087C>T; p.Pro696Leu in the same gene as shown in **Figure S1**. These probands were negative for pathogenic, likely pathogenic or VUS variants and CNVs for all genes tested by the panels, including in *BRCA1* and *BRCA2*.

All five *BRIP1* variants were re-annotated for further characterization as candidates for the study. All missense variants were predicted to be at conserved amino acid residues by all three selected in silico tools and were also predicted to be damaging at the protein level by at least one of the eight selected tools (**Table 1**). The p.Thr266Met and p. Pro696Leu were predicted to be damaging by all eight in silico tools, whereas p.Ala406Ser and p.Ser139Ala were predicted to be damaging by one or three out of the eight tools, respectively. None of the missense variants were predicted to affect splicing of *BRIP1* transcript by any of the tools.

**Table 1.**
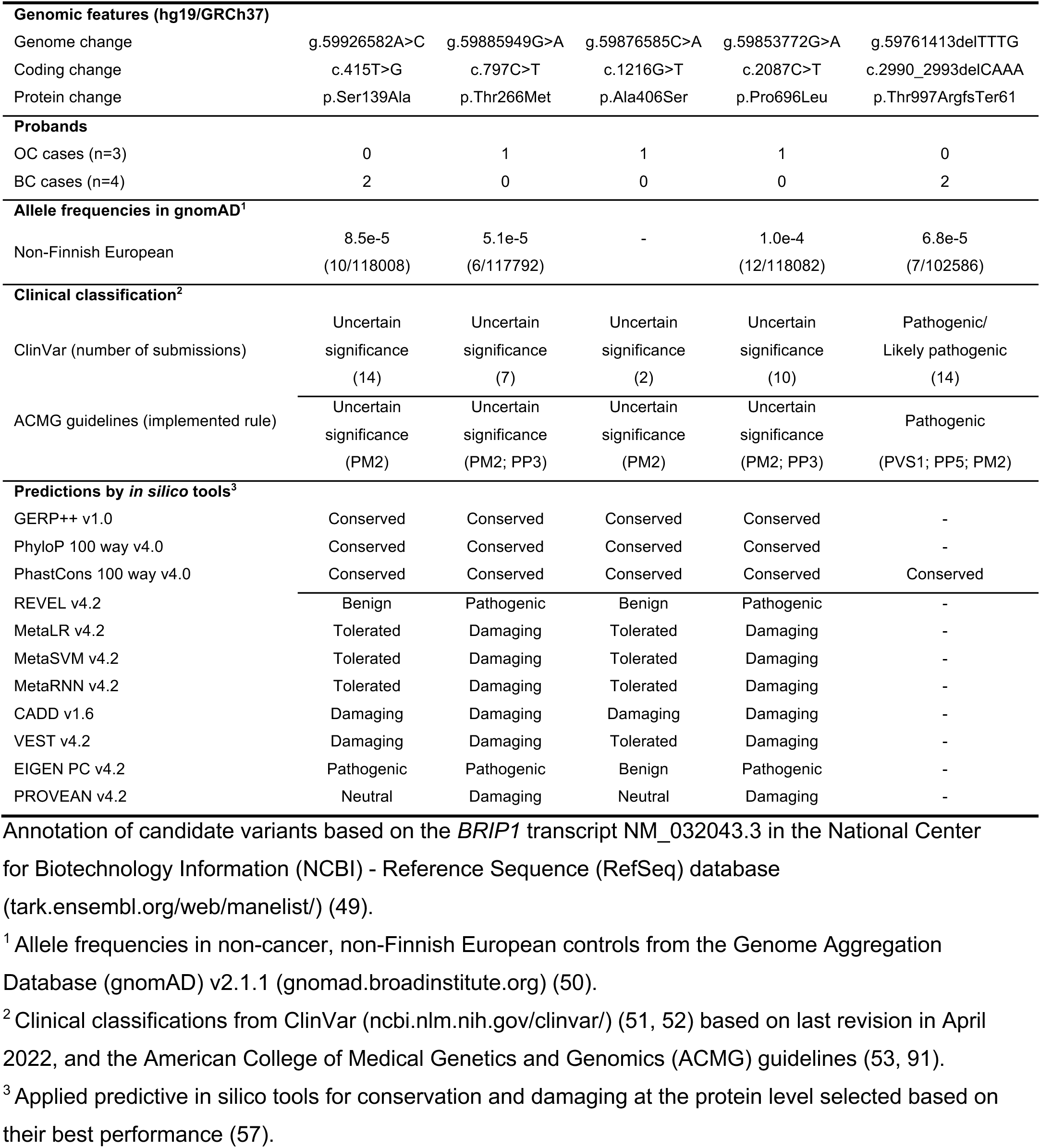
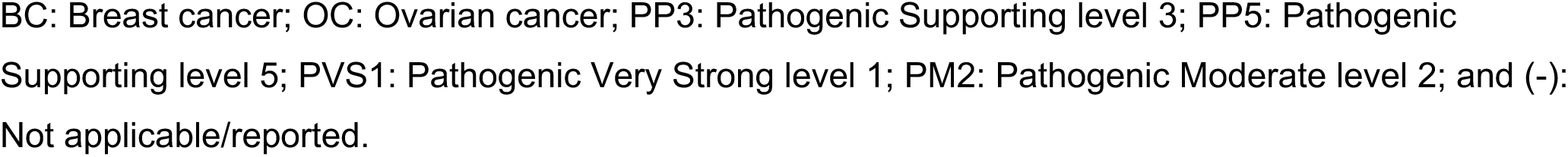
Characteristics of *BRIP1* candidate variants identified in carrier probands with ovarian or breast cancer cases of French Canadians from the adult hereditary cancer clinics.

| **Genomic features (hg19/GRCh37)** |  |  |  |  |  |
| --- | --- | --- | --- | --- | --- |
| Genome change | g.59926582A>C | g.59885949G>A | g.59876585C>A | g.59853772G>A | g.59761413delTTTG |
| Coding change | c.415T>G | c.797C>T | c.1216G>T | c.2087C>T | c.2990_2993delCAAA |
| Protein change | p.Ser139Ala | p.Thr266Met | p.Ala406Ser | p.Pro696Leu | p.Thr997ArgfsTer61 |
| **Probands** |  |  |  |  |  |
| OC cases (n=3) | 0 | 1 | 1 | 1 | 0 |
| BC cases (n=4) | 2 | 0 | 0 | 0 | 2 |
| **Allele frequencies in gnomAD<sup>1</sup>** |  |  |  |  |  |
| Non-Finnish European | 8.5e-5<br>(10/118008) | 5.1e-5<br>(6/117792) | - | 1.0e-4<br>(12/118082) | 6.8e-5<br>(7/102586) |
| **Clinical classification<sup>2</sup>** |  |  |  |  |  |
| ClinVar (number of submissions) | Uncertain<br>significance<br>(14) | Uncertain<br>significance<br>(7) | Uncertain<br>significance<br>(2) | Uncertain<br>significance<br>(10) | Pathogenic/<br>Likely pathogenic<br>(14) |
| ACMG guidelines (implemented rule) | Uncertain<br>significance<br>(PM2) | Uncertain<br>significance<br>(PM2; PP3) | Uncertain<br>significance<br>(PM2) | Uncertain<br>significance<br>(PM2; PP3) | Pathogenic<br>(PVS1; PP5; PM2) |
| **Predictions by *in silico* tools<sup>3</sup>** |  |  |  |  |  |
| GERP++ v1.0 | Conserved | Conserved | Conserved | Conserved | - |
| PhyloP 100 way v4.0 | Conserved | Conserved | Conserved | Conserved | - |
| PhastCons 100 way v4.0 | Conserved | Conserved | Conserved | Conserved | Conserved |
| REVEL v4.2 | Benign | Pathogenic | Benign | Pathogenic | - |
| MetaLR v4.2 | Tolerated | Damaging | Tolerated | Damaging | - |
| MetaSVM v4.2 | Tolerated | Damaging | Tolerated | Damaging | - |
| MetaRNN v4.2 | Tolerated | Damaging | Tolerated | Damaging | - |
| CADD v1.6 | Damaging | Damaging | Damaging | Damaging | - |
| VEST v4.2 | Damaging | Damaging | Tolerated | Damaging | - |
| EIGEN PC v4.2 | Pathogenic | Pathogenic | Benign | Pathogenic | - |
| PROVEAN v4.2 | Neutral | Damaging | Neutral | Damaging | - |

Annotation of candidate variants based on the *BRIP1* transcript NM\_032043.3 in the National Center for Biotechnology Information (NCBI) - Reference Sequence (RefSeq) database (tark.ensembl.org/web/manelist/) (49).
<sup>1</sup> Allele frequencies in non-cancer, non-Finnish European controls from the Genome Aggregation Database (gnomAD) v2.1.1 (gnomad.broadinstitute.org) (50).
<sup>2</sup> Clinical classifications from ClinVar (ncbi.nlm.nih.gov/clinvar/) (51, 52) based on last revision in April 2022, and the American College of Medical Genetics and Genomics (ACMG) guidelines (53, 91).
<sup>3</sup> Applied predictive *in silico* tools for conservation and damaging at the protein level selected based on their best performance (57).
BC: Breast cancer; OC: Ovarian cancer; PP3: Pathogenic Supporting level 3; PP5: Pathogenic Supporting level 5; PVS1: Pathogenic Very Strong level 1; PM2: Pathogenic Moderate level 2; and (-): Not applicable/reported.

As described in **Table 1**, c.2990_2993delCAAA; p.Thr997ArgfsTer61, which was reported in two probands diagnosed with BC, is rare in the cancer-free, non-Finnish European population, having a MAF of 6.8e-5, and in other populations (**Table S3**). It has been classified as pathogenic/likely pathogenic in ClinVar (Accession number: VCV000234281.23) and pathogenic by ACMG guidelines (Pathogenic Very Strong level 1 [PVS1]; Pathogenic Supporting level 5 [PP5]; and Pathogenic Moderate level 2 [PM2]). This is under the assumption that the encoded protein has a disrupted BRCT domain that would affect its interaction with BRCA1 based on a previous report that demonstrated such impact by another frameshift variant in *BRIP1* c.2992_2995del; p.Lys998GlufsTer60 (90), which is located adjacent to our variant. Our *BRIP1* frameshift variant is predicted to introduce a premature termination codon at amino acid position 61 and induce truncation of the encoding protein in the BRCT domain, if synthesized (**Figure 1-A-B**). There have been multiple reports in ClinVar of c.2990_2993delCAAA; p.Thr997ArgfsTer61 in the context of hereditary OC, BC as well as FA. The family history of this *BRIP1* frameshift variant carrier proband (PT0152) from family F1646 was consistent with features of HBC syndrome, having an early age of onset of BC (diagnosed between 30-39 years of age) and other relatives with an early age of diagnosis with BC (**Figure S1**). Whereas the same frameshift variant carrier BC proband (PT0164) from family F1656 (diagnosed between 30-39 years of age) was less indicative of harbouring features of a known hereditary cancer syndrome phenotype, though multiple types of cancer were reported in this family.

**Figure 1.**
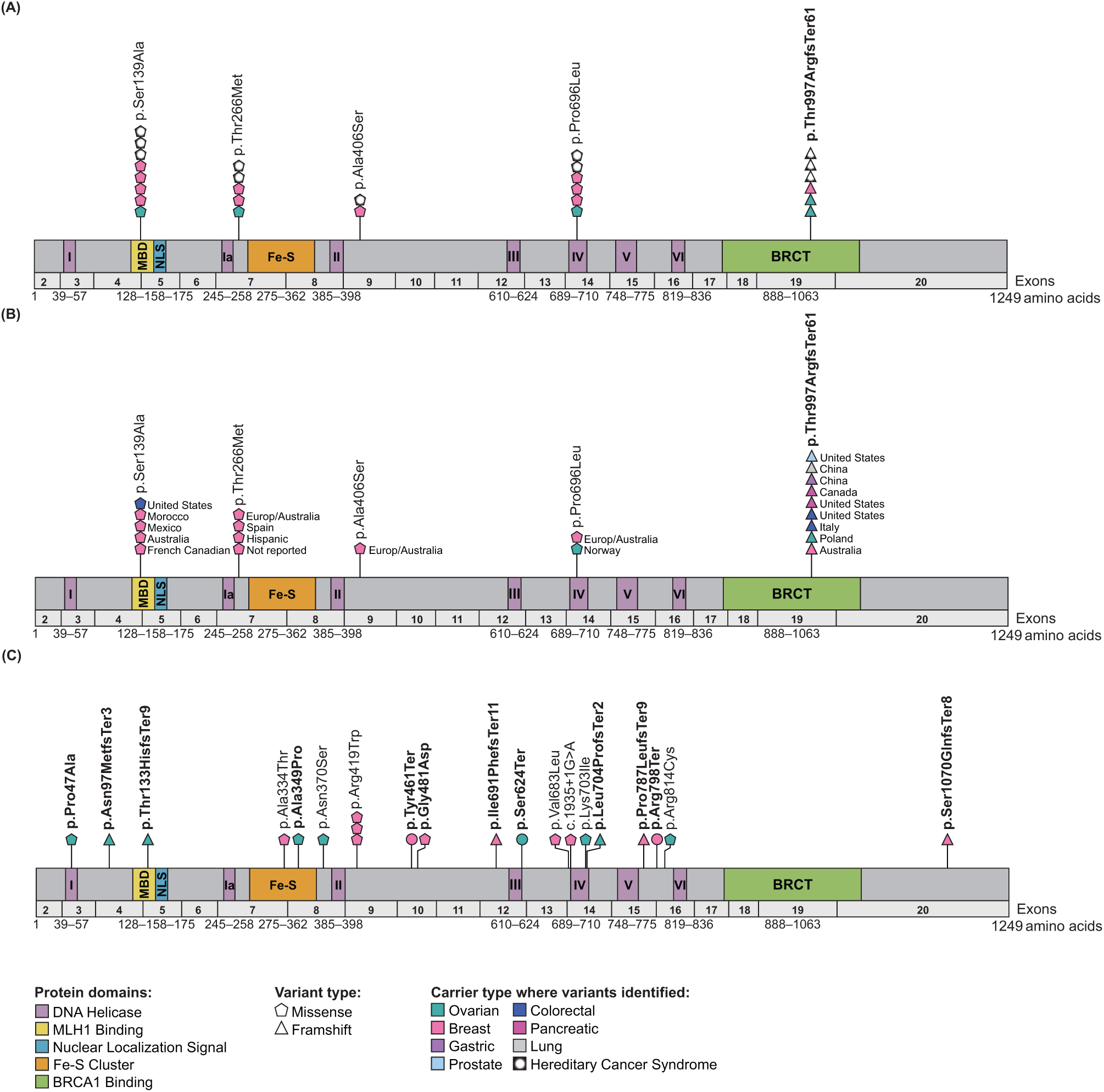
**The location of BRIP1 candidate variants in the transcript and protein domains.** Diagram of *BRIP1* transcript with protein domains indicating the location of candidate variants (see **Table S4**) identified in: (A) French Canadian ovarian (OC) and breast (BC) cancer cases in this study; different cancers reported from a review of the literature; and (C) OC and BC Pan-Cancer cases from The Cancer Genome Atlas project, mostly of European origin in this study. *BRIP1* candidate variants classified as pathogenic in ClinVar (ncbi.nlm.nih.gov/clinvar/) (51, 52) or by the American College of Medical Genetics and Genomics guidelines (53) are bolded. *BRIP1* transcript (NM_032043.3) and protein domains was based on the National Center for Biotechnology Information – Reference Sequence database (ncbi.nlm.nih.gov/protein/NP_114432.2) and (tark.ensembl.org/web/manelist/) (49).

The four *BRIP1* missense variants of interest were all rare in different populations (**Table S3**), with MAF of at least 1.0e-4 in non-Finnish European controls, or have not been identified as with c.1216G>T; p.Ala406Ser. Two probands PT0147 from family F1641 and PT0149 from family F1642 were diagnosed with BC between 40-49 and 30-39 years of age, respectively, and were reported to harbour c.415T>G; p.Ser139Ala (**Figure S1**). This variant was classified as VUS in ClinVar (VCV000132712.21) in the context of hereditary OC, BC or FA and by the ACMG Guidelines because of its rarity in the cancer-free controls (Pathogenic Moderate [PM2]). The effect of this amino acid substitution in BRIP1 is unknown though it is located in the MLH1 binding domain (MBD) (92) (**Figure 1-A-B**). Interestingly, proband PT0147 (Family F1641) reported a family history of cancer suggestive of Lynch syndrome (93), while proband PT0149 (Family F1642) reported a family history of multiple types of cancer (**Figure S1**). None of these families had a confirmed case of OC though there was a second degree relative of proband PT0147 (Family F1641) suspected of having either uterine cancer or OC. In contrast to these carriers, the probands harbouring the missense variants c.797C>T; p.Thr266Met (PT0150 from family F1636), c.1216G>T; p.Ala406Ser (PT0151 from family F1645) or c.2087C>T; p.Pro696Leu (PT0099 from family F1628) were diagnosed with OC between ages 50-59, 60-69 and 30-39 years of age, respectively (**Figure S1**). These missense variants were classified as VUS in the context of hereditary OC, BC or FA in ClinVar (VCV000128196.14; VCV000407821.7; and VCV000128167.23, respectively) and by ACMG guidelines because of their rarity in the cancer-free controls (Pathogenic Moderate Level 2 [PM2]) and in silico predictions (Pathogenic Supporting Level 3 [PP3]) (53). The effect of these amino acid substitutions in BRIP1 is also unknown though one is located in the DNA helicase domain (**Figure 1-A-B**). Though the family history of proband PT0151 (F1645) is suspicious for Lynch syndrome, with two reports of intestinal or colon cancers, the families of all the OC probands harbouring these missense variants reported multiple cancer types. There were no striking characteristics of the family history of cancer in probands PT0150 (Family F1636) and PT0099 (Family F1628) (**Figure S1**).

### Multiple carriers of candidate *BRIP1* variants were identified in defined FC cancer study groups

The carrier frequencies of *BRIP1* candidate variants were determined in FC study groups comprised of familial and sporadic OC or BC cases, regardless of their status of *BRCA1* and *BRCA2* pathogenic variants and in population-matched controls. We did not identify any other carriers of c.2990_2993delCAAA in any of our study groups. However, we identified carriers of each of our missense variants in at least one of our FC study groups (**Table 2**). We determined that the frequency of cancer carriers ranged from 0-0.7% versus 0-0.2% in the population-matched controls, depending on the variant and the group investigated (**Table 2**). However, the carrier frequency of each variant in a defined cancer group was not significantly different to that of the control group. It is interesting to observe that carriers of missense variants were mostly identified in sporadic BC cases in contrast to OC cases.

**Table 2.** Carrier frequencies of candidate *BRIP1* variants in French Canadian cancer cases and population-matched controls.

| *BRIP1* variant | Study groups<sup>1</sup> | Cancer cases tested<sup>1</sup> | Number of tested participants (or families) per study group | Number of variant carriers (%) | p-value<sup>2</sup> |
| --- | --- | --- | --- | --- | --- |
| c.415T>G;<br>p.Ser139Ala | OC families | OC | 66 (47) | 0 | - |
|  | HBOC families | OC or BC | 49 (49) | 0 | - |
|  | HBC families | BC | 142 (142) | 0 | - |
|  | Sporadic OC cases | OC | 435 | 0 | - |
|  | Sporadic BC cases | BC | 563 | 1/563 (0.2) | 1.000 |
|  | FC sequencing-based controls | - | 1025 | **2/1025 (0.2)** | - |
| c.797C>T;<br>p.Thr266Met | OC families | OC | 66 (47) | 0 | - |
|  | HBOC families | OC or BC | 49 (49) | 0 | - |
|  | HBC families | BC | 142 (142) | **1/142 (0.7)** | 0.122 |
|  | Sporadic OC cases | OC | 435 | **1/435 (0.2)** | 0.298 |
|  | Sporadic BC cases | BC | 563 | **2/563 (0.4)** | 0.126 |
|  | FC sequencing-based controls | - | 1025 | 0 | - |
| c.1216G>T;<br>p.Ala406Ser | OC families | OC | 66 (47) | 0 | - |
|  | HBOC families | OC or BC | 49 (49) | 0 | - |
|  | HBC families | BC | 142 (142) | 0 | - |
|  | Sporadic OC cases | OC | 435 | 0 | - |
|  | Sporadic BC cases | BC | 563 | **2/563 (0.4)** | 0.126 |
|  | FC sequencing-based controls | - | 1025 | 0 | - |
| c.2087C>T;<br>p.Pro696Leu | OC families | OC | 66 (47) | 0 | - |
|  | HBOC families | OC or BC | 49 (49) | 0 | - |
|  | HBC families | BC | 142 (142) | 0 | - |
|  | Sporadic OC cases | OC | 435 | 0 | - |
|  | Sporadic BC cases | BC | 563 | **1/563 (0.2)** | 0.355 |
|  | FC sequencing-based controls | - | 1025 | 0 | - |
| c.2990_2993delCAAA;<br>p.Thr997ArgfsTer61 | OC families | OC | 66 (47) | 0 | - |
|  | HBOC families | OC or BC | 49 (49) | 0 | - |
|  | HBC families | BC | 142 (142) | 0 | - |
|  | Sporadic OC cases | OC | 435 | 0 | - |
|  | Sporadic BC cases | BC | 563 | 0 | - |
|  | FC sequencing-based controls | - | 1025 | 0 | - |

<sup>1</sup> Study groups comprised of ovarian cancer (OC), breast cancer (BC), hereditary breast and ovarian cancer (HBOC) syndrome and hereditary breast cancer (HBC) syndrome families, all from the French Canadian (FC) population of Quebec.
<sup>2</sup>Two-tailed *p*-values (not adjusted for multiple testing) calculated using Fisher's exact test in pair-wise comparisons between variant carriers of each cancer study group and population-matched controls. Dash (-) denotes not applicable.

It was not possible to determine the carrier frequency of any of the *BRIP1* candidate variants in 8493 genotyping-based FC controls (**Table S1**) as none of these variants were represented on any of these genotyping arrays. However, we were able to impute c.797C>T as this variant was present in the HRC.r1 haplotype reference panel; no carriers were identified among the genotyping-based controls, suggesting that this variant is rare in the cancer-free FC population.

With our expectation that some of our candidates may occur with a higher frequency in the FC population due to genetic drift (3), we investigated our candidate *BRIP1* variants in additional cancer cases. We genotyped PBL DNA or surveyed available genetic data of 534 additional OC and 52 sporadic early-onset OC cases for carriers of our candidate *BRIP1* variants. We identified four carriers among the additional group of OC cases, and none in the early-onset cases, harbouring c.415T>G; p.Ser139Ala (PT0120), c.1216G>T; p.Ala406Ser (PT0200), c.2087C>T; p.Pro696Leu (PT0119) or c.2990_2993delCAAA; p.Thr997ArgfsTer6 (PT0156) (**Table S4**). The carrier frequency of each variant in these study groups was less than 2%, which is consistent with the low carrier frequency observed for these variants in the other defined FC study groups (**Table 2**).

### Genetic analyses of BC and OC cases and controls not selected for FC ancestry identified candidate *BRIP1* variants

Using our criteria for identifying clinically relevant candidate *BRIP1* variants, we investigated genetic data from the germline of 412 OC and 1072 BC Pan-Cancer – TCGA cases and 134,187 cancer-free, non-Finnish European controls. We identified carriers of nine variants in 8/412 (1.9%) OC cases and ten variants in 9/1076 (0.8%) BC cases (**Figure 1-C** and **Table S4**), which included one BC carrier of c.415T>G; p.Ser139Ala, a candidate variant that was also identified in our FC cases (**Table 1).** These variants were identified in 0.001-0.09% of the non-cancer, non-Finnish European controls in gnomAD (**Table S4**). There were ten LoF (three nonsense, six frameshift and one canonical alternative splicing) and nine missense variants, including c.1109A>G; p.Asn370Ser that was predicted by SpliceAI (66) to affect splicing that may result in donor loss (**Table S4**). Eight of these variants were located in biologically relevant domains of BRIP1 comprised of the MBD and iron-sulfur (Fe-S) (94) and one of the DNA helicase motifs (32) (**Figure 1-C**).

### In cellulo assays revealed deleterious *BRIP1* candidate variants affected cellular sensitivity to cisplatin but not to olaparib or talazoparib

To explore the functionality of BRIP1 protein encoded by candidate variants identified in our FC study groups, we generated stable cell lines expressing: p.Ser139Ala, p.Thr266Met, p.Ala406Ser, p.Pro696Leu, p.Thr997ArgfsTer61, a BRIP1 WT and an empty vector (EV) using the AAVS1 genomic editing system in BRIP1-depleted cells (**Figure 2A-B**) (85). Two *BRIP1* variants were included as controls: c.1045G>C; p.Ala349Pro (27, 28), which is classified in ClinVar as pathogenic/likely pathogenic (VCV000030535.14) or as a VUS by ACMG (Pathogenic Moderate level 2 [PM2]) and c.2220G>T; p.Gln740His (27, 28), which is classified in ClinVar as likely benign (VCV000133752.34) or as a VUS by ACMG (Pathogenic Moderate level 2 [PM2]). BRIP1 p.Ala349Pro was selected as a positive control which was predicted to be damaging at the protein level by six out of our eight selected in silico tools (38, 56); and BRIP1 p.Gln740His was selected as a negative control which was predicted to be damaging by only one out of the eight predictive tools.

**Figure 2.**
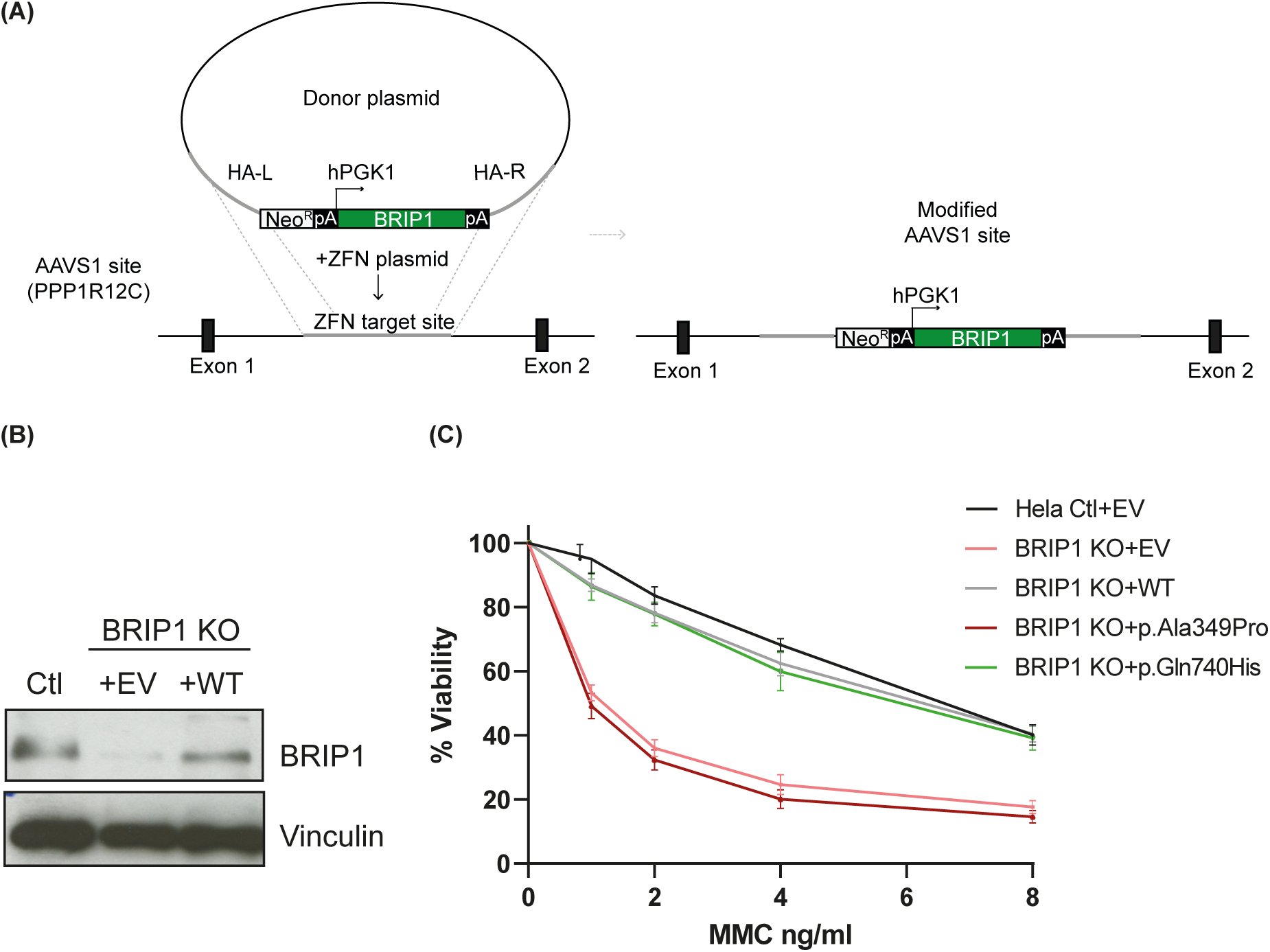
**Strategy used to assess the functional impact of BRIP1 variants in a mammalian cell system**. (A) Scheme representing the AAVS1 genomic editing system used to complement BRIP1 knock-out (KO) cells. (B) Hela control (Ctl) cells and BRIP1 KO cells completed with empty vector (EV) or BRIP1 wild-type (WT) using the AAVS1 system were confirmed by western blot analyses of BRIP1 protein expression, and vinculin was used as a loading control. (C) Survival curves of Hela BRIP1 cells stably complemented with constructs of BRIP1 WT, BRIP1 p.Ala349Pro (a likely pathogenic/pathogenic variant as a control), BRIP1 p.Gln740His (a likely benign variant as a control), or EV and plated in triplicate in a 96 well plate. Cell viability was monitored following mitomycin (MMC) treatment for 96 hours and was assessed by counting remaining nuclei. Experiments were performed in three biological replicates.

BRIP1 activity is critical for mediating the repair of DNA ICLs, and cells deficient for this gene are sensitive to ICL-inducing agents such as MMC and cisplatin (92, 95). Given this phenotype, we assessed the sensitivity of the cells containing the interrogated BRIP1 variants to increasing concentrations of either MMC or cisplatin. As expected, Hela BRIP1 KO cells complemented with the EV were more sensitive to both MMC and cisplatin when compared to the Hela control cells (Ctl) (**Figure 2C** and **Figure S2**) (27). Complementation with the BRIP1 WT rescued the cells sensitivity from the effect of ICL-inducing agents. This phenotype was also observed in BRIP1 KO cells complemented with BRIP1 p.Gln740His, the negative control. However, cells complemented with BRIP1 p.Ala349Pro, the pathogenic/likely pathogenic control, failed to confer resistance to MMC or cisplatin (**Figure 2C** and **Figure S2**). A similar profile was observed in cells complemented with the EV. As previously demonstrated (27), greater sensitivity to MMC and cisplatin was also observed in BRIP1-depleted U2OS cells complemented with the EV or BRIP1 p.Ala349Pro, and resistance was partially recovered with the BRIP1 WT (**Figure S3**).

We next determined whether BRIP1-depleted cells expressing the selected BRIP1 variants were able to confer resistance to the ICL-inducing agents MMC and cisplatin (**Figure 3** and **Figure S2**).

**Figure 3.**
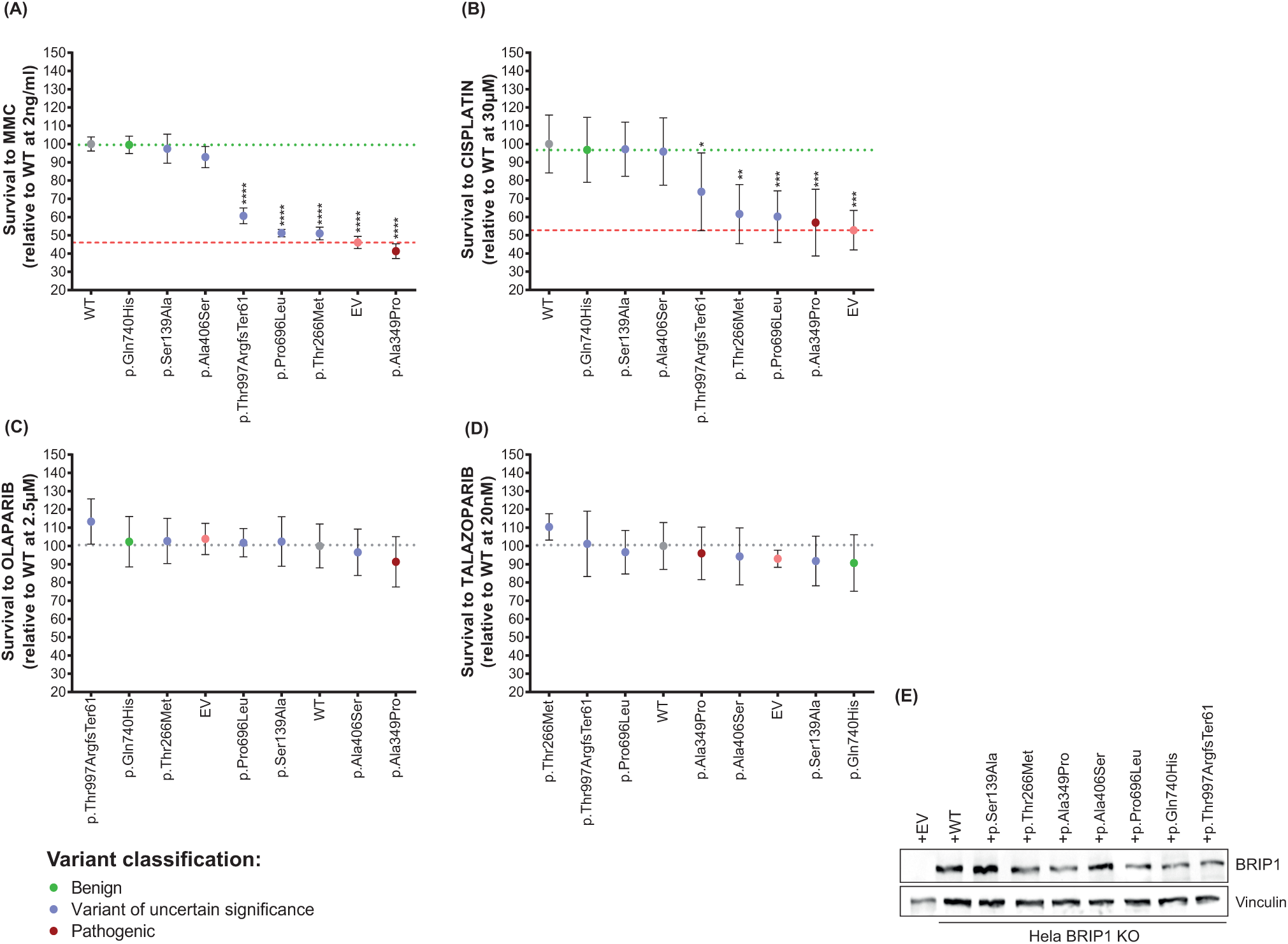
**Drug sensitivity of cells expressing BRIP1 variants to DNA inter– and intra-strand crosslinks inducing agents and poly-ADP-ribose polymerase inhibitors**. Sensitivity of BRIP1 knock-out (KO) cells stably complemented with constructs of BRIP1 variants or empty vector (EV) to the DNA inter– or intra-strand crosslinks (ICLs) inducing agents: (A) Mitomycin C (MMC) and (B) cisplatin; and to poly-ADP-ribose polymerase (PARP) inhibitors (C) olaparib and (D) talazoparib. Sensitivity profiles were determined with the BRIP1 wild-type (WT) set as 100% at the concentrations of 2ng/ml for MMC (A), 30μM for cisplatin (B), 2.5μM of olaparib (C) and 20nM for talazoparib (D). Survival data from each BRIP1 variant are sorted in descending order in response to drug sensitivity and presented based on the mean (±standard error of the mean [SEM]) from at least 3 independent experiments, performed in triplicate. Statistical significance was determined by One-way ANOVA test with Dunnett’s multiple comparison post-test. (*) *P <* 0.05; (**) *P <* 0.01; (***) *P <* 0.001 and (****) *P <* 0.0001. (E) Western blot analysis showing BRIP1 protein expression in BRIP1-depleted cells after stable complementation with the indicated variants, with vinculin as a loading control.

BRIP1 p.Ser139Ala and p.Ala406Ser behaved similarly to the BRIP1 WT in terms of the ability to rescue the viability of BRIP1-depleted cells to MMC and to cisplatin, indicating that these missense variants do not impact the functionality of BRIP1 to resolve ICLs (**Figure 3A-B**). However, cells expressing BRIP1 p.Thr266Met, p.Pro696Leu or p.Thr997ArgfsTer61 were unable to rescue the sensitivity of BRIP1-depleted cells. BRIP1 p.Thr266Met and p.Pro696Leu stood out with the highest sensitivity to the ICL-inducing drugs (**Figure 3A-B**), with survival percentages each of 51% relative to the WT at dose of 2ng/ml of MMC and 61% and 60% relative to WT at a dose of 30μM of cisplatin, respectively. Using the same criteria, the relative survival of EV was 46% to MMC and 52% to cisplatin, BRIP1 p.Ala349Pro was 41% to MMC and 56% to cisplatin, while BRIP1 p.Thr997ArgfsTer61 sensitivity was 60% and 73% to MMC and cisplatin, respectively (**Figure 3A-B**). These results provide evidence in favor of the possible impaired function of BRIP1 p.Thr266Met, p.Pro696Leu and p.Thr997ArgfsTer61 variants. Survival curves for all variants are depicted in **Figure S2**. For further validation, variants were also tested in the U2OS BRIP1-depleted cell line (**Figure S3**). Similar results to those obtained in Hela cells were observed. However, the dynamic range between U2OS BRIP1-depleted cells complemented with the EV and the WT was proportionately smaller relative to Hela cells. In U2OS cells, BRIP1 p.Ala406Ser appears to have a partial complementation (**Figure S3C** and **I**). Concerning protein expression, the BRIP1 variants tested here lead to a protein product as detected by immunoblotting in our experimental Hela and U2OS cell line models (**Figure 3E**, **Figure S2K** and **Figure S3K**). Considering the impact of PARP inhibitors on the clinical management of BC and OC patients (96), cells expressing our BRIP1 variants were also tested for the sensitivity to PARP inhibitors, olaparib or talazoparib. As previously demonstrated (97), cells depleted in BRIP1 have no greater sensitivity to either olaparib or talazoparib. Complementation with the BRIP1 WT or any of our BRIP1 variants did not alter the resistance profile to either PARP inhibitor (**Figure 3C-D** and **Figure S4**).

## DISCUSSION

We investigated five rare *BRIP1* variants that were initially reported in OC or BC cases by the adult hereditary cancer clinics for candidacy as clinically relevant variants. Our genetic analyses of these variants: (1) assessed bioinformatically their potential impact on gene function; (2) investigated their carrier frequency in defined cancer study groups comprised of familial and sporadic OC and BC cases and population-matched controls from a population exhibiting genetic drift; and (3) assessed bioinformatically other candidate variants in *BRIP1* and investigated their carrier frequency in ancestrally diverse cancer study groups and controls. We also assayed biologically, the impact of these five *BRIP1* variants on the encoded protein function based on the current known role of BRIP1 in DNA repair (17) and in cellulo sensitivity to chemotherapeutic agents such as MMC and cisplatin and targeted therapeutic agents such as PARP inhibitors (98, 99). Collectively, our findings from these assays suggest that a frameshift variant c.2990_2993del; p.Thr997ArgfsTer6 and two of the four missense variants c.797C>T; p.Thr266Met and c.2087C>T; p.Pro696Leu likely affect BRIP1 function.

The identification of multiple carriers of each of our *BRIP1* candidate variants is likely attributable to shared ancestry of the FC population of Quebec (35, 38, 39, 100). We could not determine whether there was a shared genome segment identical-by-descent due to the paucity of carriers of each variant though we have been able to demonstrate shared ancestry using haplotype analyses of the carriers of most frequently occurring pathogenic variants in *BRCA1*, *BRCA2* (35, 100, 101), *PALB2* (37), *RAD51C* (38) and *RAD51D* (38, 39) in the context of OC and BC in the FC population (3). In 2008, an early independent study of *BRIP1* in FC BC cases from HBC or HBOC families of Quebec reported 42 variants in *BRIP1* but concluded that none are likely clinically relevant (41). We reassessed these variants with our selected in silico tools and retrieved current information from genetic databases (see **Table S6**), and indeed, none were predicted to be biologically relevant. The only variant found in common with our study of the FC cancer cases was c.415T>G; p.Ser139Ala, but we showed that this variant is unlikely to affect the protein function. We classified 86% of these variants as benign or likely benign based on reports in ClinVar and/or by ACMG guidelines, which is not surprising as 50% of variants are common having MAF>0.01 in the FC controls. The genetic heterogeneity observed in *BRIP1* variant carriers is consistent with the germline genetic landscape of the FC population of Quebec (3). The differences in carrier frequencies of our variants in *BRIP1* as well as those observed in *BRCA1*, *BRCA2, PALB2, RAD51C* and *RAD51D* are expected in FCs and consistent with the genetic drift that has been attributed to the waves of localized expansion of this population that occurred in Quebec since 1608 (102–104). Given the European ancestry of FCs, it is not surprising that all five candidate *BRIP1* variants were also identified in the germline of cancer cases in the literature (see **Figure 1-B**). Moreover, the overall low carrier frequency of candidate *BRIP1* variants in FC cancer study groups and the Pan-Cancer – TCGA cases is consistent with the overall low carrier frequency (approximately <2%) of pathogenic *BRIP1* variants that have been reported in cancer cases from other populations (9, 15).

Although the role of our candidate *BRIP1* variants in conferring risk to OC and BC remains to be determined, our in cellulo analyses suggest that some affect BRIP1 function. BRIP1 binds directly to BRCA1 via BRCT motifs which play a critical role in BRCA1 stability to mediate repair of double-stranded DNA breaks (14, 17, 105). Based on our current knowledge, pathogenic variants in the BRCT domain of BRIP1 negatively affected the repair of double-stranded DNA breaks by abrogating the BRIP1-BRCA1 interaction (23) rendering cells sensitive to cisplatin (27, 28, 32). We showed in our in cellulo assays, that our BRIP1 variant p.Thr997ArgfsTer61 predicted to affect the BRCT domain, though expressed impaired cellular sensitivity to MMC and cisplatin. Though speculative, the ability of BRIP1 p.Thr997ArgfsTer61 to interact with BRCA1 may have been impaired as a consequence of the loss of an intact BRCT domain for BRCA1 binding. Another frameshift variant in *BRIP1* c.2992_2995del; p.Lys998GlufsTer60 which affects an adjacent amino acid has also been shown to be expressed in cells (90), suggesting that transcripts from these variants may not elicit nonsense mediated decay. Our functional assessment determined that candidate variants p.Thr266Met, p.Pro696Leu and p.Thr997ArgfsTer61 exhibited loss of BRIP1 WT function upon exposure to MMC and cisplatin, while p.Ser139Ala and p.Ala406Ser did not alter cellular sensitivity to these ICL-inducing agents. The proximity of p.Thr266Met and p.Pro696Leu to any one of the helicase domains in BRIP1 may account for effect on the protein function (27, 28) in our assays and warrants further biochemical characterization of helicase activity. Though lack of BRIP1 results in HR deficiency and loss of replication fork protection, it does not result in PARP inhibitor-induced single-stranded DNA breaks (97). Thus, none of the five variants expressing cells exhibited sensitivity to PARP inhibitors, consistent with independent reports of response to WT and variant BRIP1 (97, 106). This may have clinical implications for the management of OC and BC patients who are carriers of pathogenic *BRIP1* variants (107, 108) and (nccn.org/guidelines/category_2). Indeed, it has been shown that BC tumour DNA from *BRIP1* carriers did not exhibit a mutational signature characteristic of HR defects a signature exhibited in *BRCA1* and *BRCA2* carriers exhibiting sensitivity to PARP inhibitors (109). Although we were able to cultivate BRIP1-deficient cell lines as also reported by other groups, we had considerable difficulty performing complementation of small interfering RNA (siRNA) BRIP1-deficient cells with a WT construct using a transient transfection system (28). To overcome this issue, we generated stable cell lines using the AAVS1 system in a CRISPR Cas9 KO background that was able to rescue BRIP1 WT protein. Genomic editing using a donor guide containing the studied variants could be applied to further overcome this barrier.

The bioinformatic tools selected to predict the effect our missense candidate variants on protein function align in part with the results of our MCC and cisplatin sensitivity assays. It is notable that six out of the eight in silico tools, selected for their best performance (57, 60), predicted that p.Thr266Met and p.Pro696Leu to be damaging. In contrast, only one in silico tool predicted that p.Ala406Ser and p.Ser139Ala to be damaging, and two other tools also predicted that p.Ala406Ser to be damaging. The prediction scores of the positive and negative controls, p.Ala349Pro and p.Gln740His (27, 28), which are classified as likely pathogenic and benign, respectively in the ClinVar Database and by ACMG guidelines, were consistent with our expectation of these variants as positive and negative controls in our assays. Our observations highlight the relevance of performing functional assays on missense variants, when possible, though this may not be feasible in medical genetics settings.

Due to the small number of carriers in cancer cases, particularly in familial cases, this study was underpowered to address differences in *BRIP1* carrier frequencies in OC versus BC cases in our FC population. Moreover, it was not feasible to screen all the FC cancer cases investigated in this study for *BRIP1* variants. Nonetheless, there were more carriers of c.797C>T; p. Thr266Met and c.2087C>T; p.Pro696Leu in sporadic BC cases (5/563, 0.9%) versus sporadic OC cases (1/435, 0.2%) though this was not statistically different. It was not unexpected to find a carrier among our HBC families (1/142; 0.7%), given the fact that *BRIP1* was originally reported as a BC predisposing genes by investigating HBC families (1). One of the first reports investigating the germline of selected candidate genes involved in the HR pathway in sporadic OC cases reported four carriers of *BRIP1* pathogenic variants, two with a family history of BC (110). A recent population-based study investigating genes involved in BC risk reported a statistical difference in carriers of pathogenic *BRIP1* variants in cases with a family history of BC versus controls (20/6361 [0.31%]; odds ratio= 2.15; 95% Confidence Interval [CI]: 1.25 to 3.58); p=0.004) (12), a result consistent with another study (9) and the original report describing pathogenic variant*s* in *BRIP1* in familial BC cases (1).

A literature review of our candidate *BRIP1* variants, revealed that one of our likely pathogenic variants, c.2990_2993delCAAA; p.Thr997ArgfsTer61, occurred in the context of hereditary cancers other than BC or OC, such as colorectal and prostate cancers (110) (see **Figure 1-B**). Interestingly, the same report described three carriers of *BRIP1* pathogenic variants each having a family history of colorectal cancer or uterine cancer (110). Carriers of pathogenic variants in *BRIP1* have been reported in colorectal cancer cases with a family history of colorectal cancer and other cancer types, or early-onset disease (93, 111–116), that were not explained by known colorectal cancer predisposing genes (117). Pathogenic variants in *BRIP1* have also been reported in familial and/or early-onset prostate cancer cases not explained by known prostate cancer risk genes (118–122). These findings suggest that harbouring pathogenic *BRIP1* variants increases risk for developing colorectal (123) or prostate cancer (118). Thus, pathogenic *BRIP1* variants may also be involved in conferring risk to a variety of cancers other than OC or BC, though penetrance has yet to be determined for these other types of cancer.

In conclusion, we applied a strategy to characterize candidate *BRIP1* variants in BC and OC cases that were initially identified in medical genetics settings, providing evidence for their role in hereditary and sporadic disease in a defined population exhibiting genetic drift and inferred their biological impact applying in cellulo assays. As we have demonstrated in previous studies of other known BC and OC predisposing genes (38), our strategy in investigating the germline of the genetically unique FC population of Quebec has the potential of identifying variants in cancer predisposing genes that may also be relevant to other populations. Our in cellulo assays involving response to cisplatin and PARP inhibitors revealed the potential impact in abrogating protein function for some of the variants, providing insights on their clinical implications that warrant further investigation in patients harbouring *BRIP1* variants. Although penetrance for *BRIP1* variants identified in the FC population has yet to be established, collectively, our findings further support the classification of c.2990_2993del; p.Thr997ArgfsTer6 as pathogenic, and provides evidence for the reclassification of c.797C>T; p. Thr266Met and c.2087C>T; p.Pro696Leu from missense variants of uncertain significance to likely pathogenic.

## SUPPLEMENTARY DATA

Supplementary Data is available in:

**Table S1.** Description of different study groups investigated in this study; **Table S2.** List of customized primers for site-directed mutagenesis; **Table S3.** Frequencies of candidate variants identified in French Canadians with ovarian or breast cancers in different cancer-free populations in the Aggregation Genome Database; **Table S4.** All *BRIP1* candidate variants identified in ovarian or breast cancer cases in this study; **Table S5.** Clinico-pathological characteristics of ovarian or breast cancer cases carrying one of the *BRIP1* candidate variants; **Table S6.** All *BRIP1* candidate variants identified in familial breast cancer cases and controls of French Canadians by Guénard et al., 2008 (41); **Figure S1.** Pedigrees of index ovarian or breast cancer cases carrying a candidate *BRIP1* variant reported by the adult hereditary cancer clinics; **Figure S2**. Sensitivity curves of BRIP1 variants to DNA inter– or intra-strand crosslinks inducing agents in Hela cells; **Figure S3.** Sensitivity curves of BRIP1 variants to DNA inter– or intra-strand crosslinks inducing agents in U2OS cells; and **Figure S4.** Sensitivity curves of BRIP1 variants to poly (ADP-ribose) polymerase inhibitors in Hela cells

## Data Availability

All data produced in the present study are available upon reasonable request to the authors

## ACKNOWLEDGEMENTS

We acknowledge Patrice Peron and Marie-France Hivert who established the Gen3G cohort and sequencing project, and Pierre-Étienne Jacques and Simon Gravel who contributed to the generation and analysis of sequencing data in Gen3G project. We thank Manon De Ladurantaye and Lise Portelance for providing clinical data and DNA samples from the RRCancer biobank; and Supriya Behl, Nancy Hamel and Celine Domecq for providing genetic data for sporadic breast cancer cases. We thank Sharon Cantor for the gift of BRIP1 knock-out cell lines.

## AUTHOR CONTRIBUTIONS

Conceived and designed project: L.M., W.M.A., P.N.T. and J-Y.M; Conducted experiments: L.M. and W.M.A.; Analyzed data: L.M., W.M.A., T.R., K.K.O., C.T.F., C.S., P.N.T. and J-Y.M; Provided samples and/or data: A-M.M-M., D.P., W.D.F., Z.E.H., D.S., G.R. J.B., T.P. and L.B.; Statistical supervision: C.M.T.G.; Genetic and bioinformatics supervision: P.N.T. and J.R.; Project supervision: P.N.T. and J-Y.M. Initial manuscript preparation: W.M.A. and L.M.; Manuscript editing: All.

## ETHICS APPROVAL

This project has received approval from The McGill University Health Centre (MUHC) REB (MP-37–2019-4783).

