## Supplemental Figures S1-S4 for "Genetic and molecular analyses of candidate germline *BRIP1/FANCJ* variants implicated in breast and ovarian cancer"

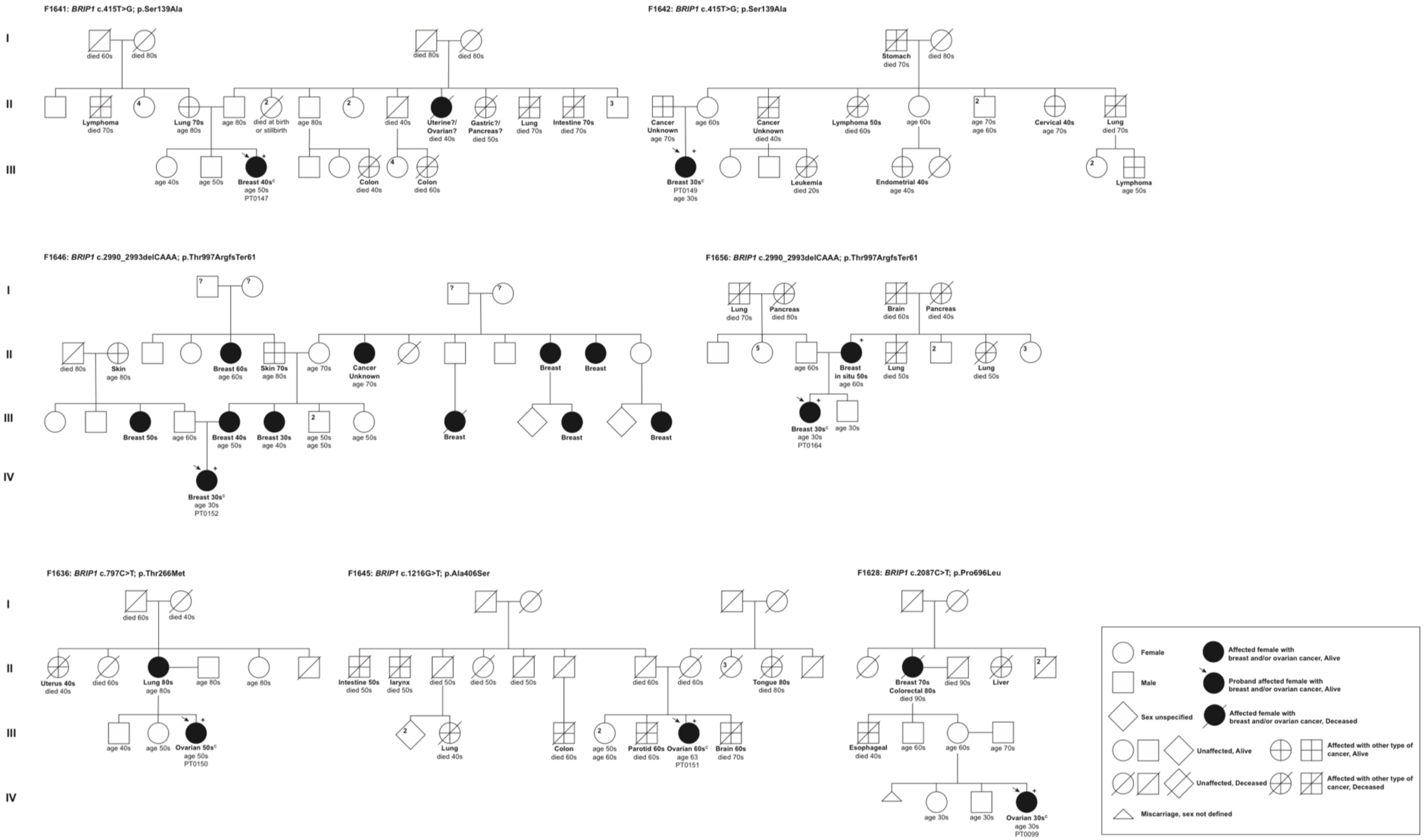


**Figure S1.** Anonymized pedigrees of index ovarian or breast cancer cases carrying a candidate *BRIP1* variant reported by the adult hereditary cancer clinics.

Carrier status of index cases (arrow) tested positive are denoted by plus sign. All carriers were found in a heterozygous state. Age ranges in years is shown with cancer diagnosis or death. Unconfirmed cancer status as reported by the index case was denoted by a question mark (?) beside the reported cancer. All breast cancer (BC) cases were invasive unless stated otherwise (see **Table S5**). Superscript C denotes histological subtype that was confirmed by pathology report.

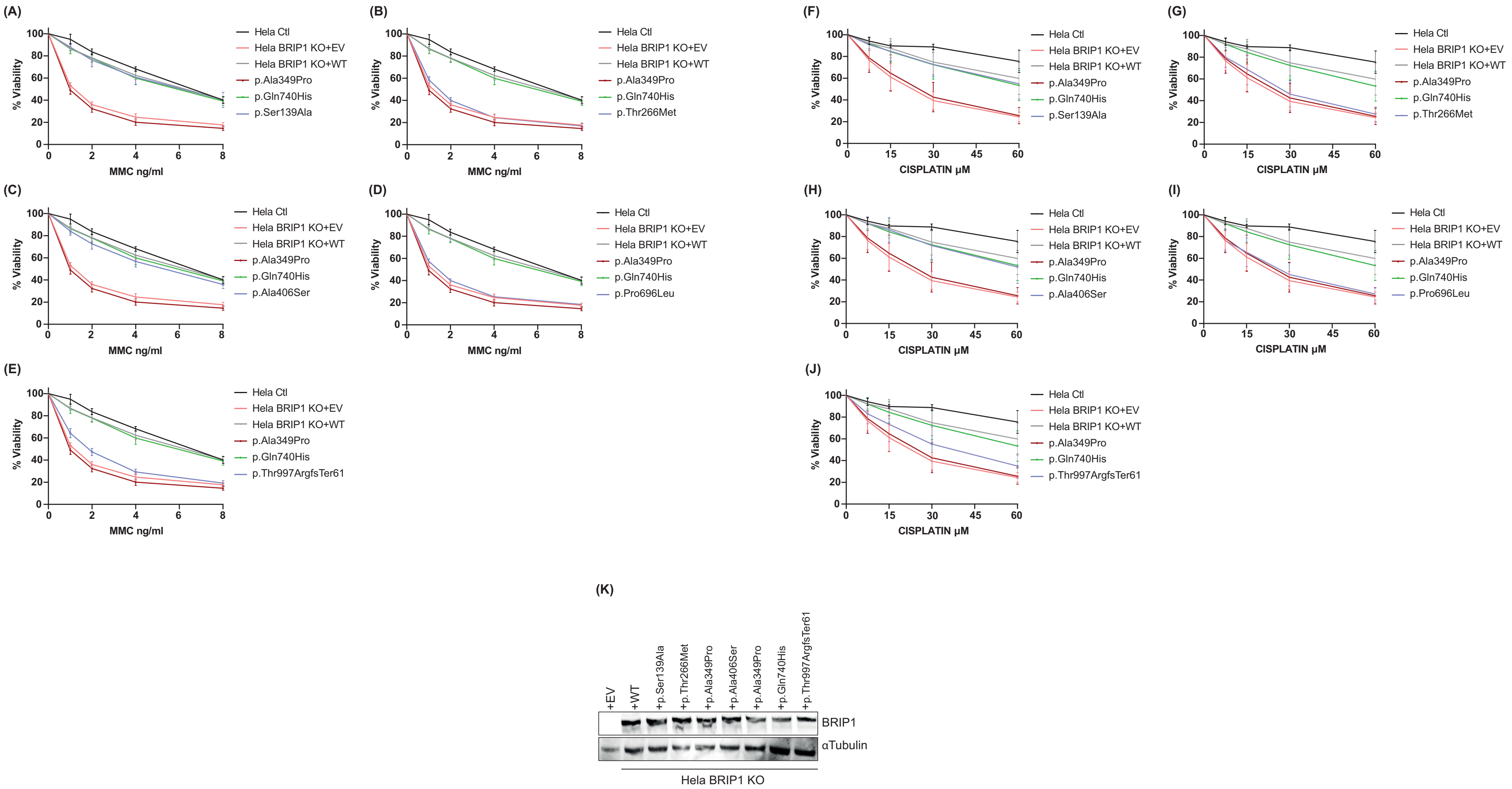


**Figure S2.** Sensitivity curves of BRIP1 variants to DNA inter- or intra-strand crosslinks inducing agents in Hela cells.
Survival curves contrasting the abilities of BRIP1 wild-type (WT) and the indicated variants, including the empty vector (EV), to rescue mitomycin C (MMC) (A-E) and cisplatin (F-J) resistance in Hela BRIP1-depleted cells. (K) Western blot representing expression of the indicated variants in Hela BRIP1 depleted cells.

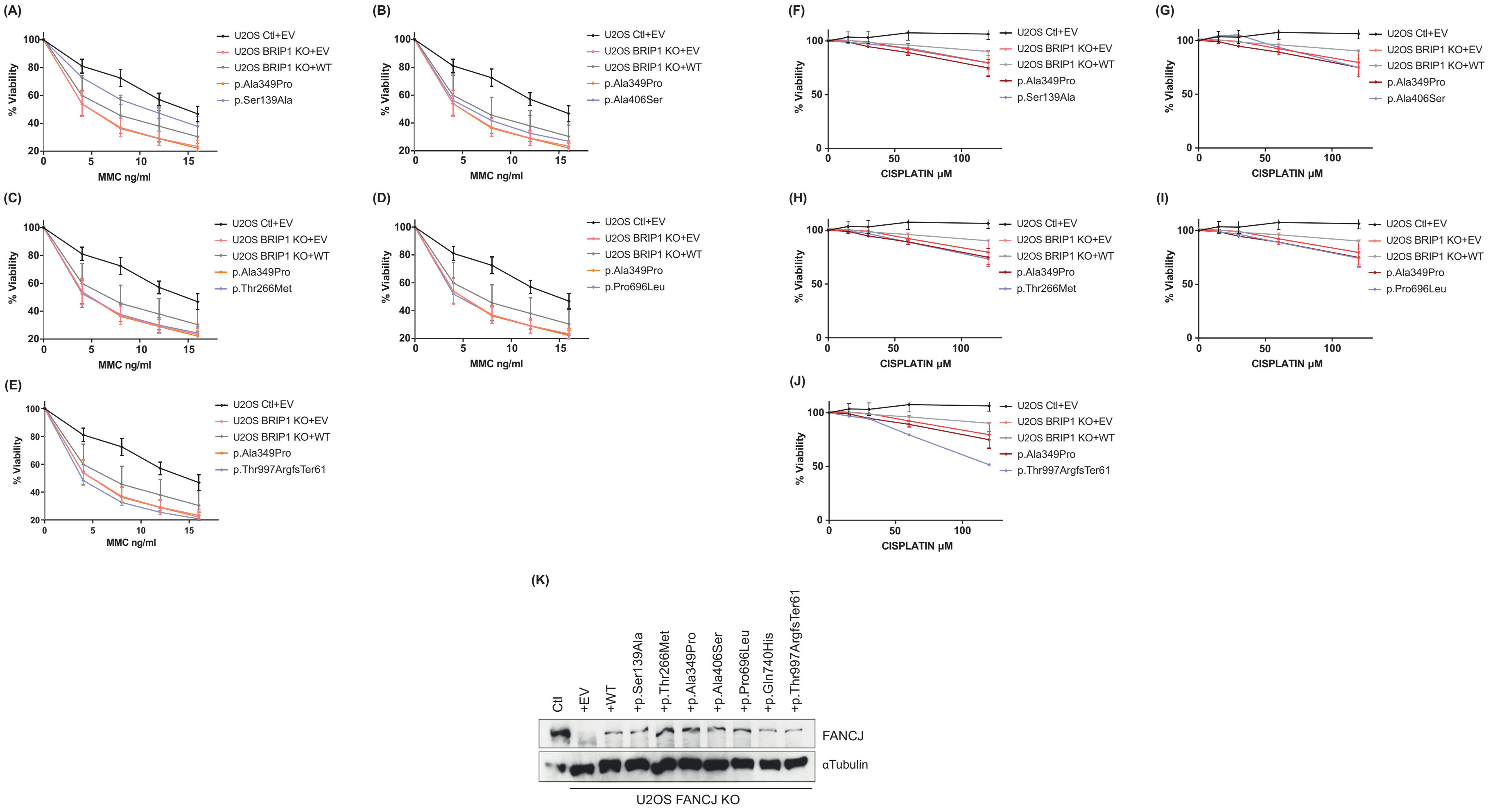


**Figure S3.** Sensitivity curves of BRIP1 variants to DNA inter- or intra-strand crosslinks inducing agents in U2OS cells.
Survival curves contrasting the abilities of BRIP1 wild-type (WT) and the indicated variants, including the empty vector (EV), to rescue mitomycin C (MMC) (A-E) and cisplatin (F-J) resistance in U2OS BRIP1-depleted cells. (K) Western blot representing expression of the indicated variants in U2OS BRIP1 depleted cells.

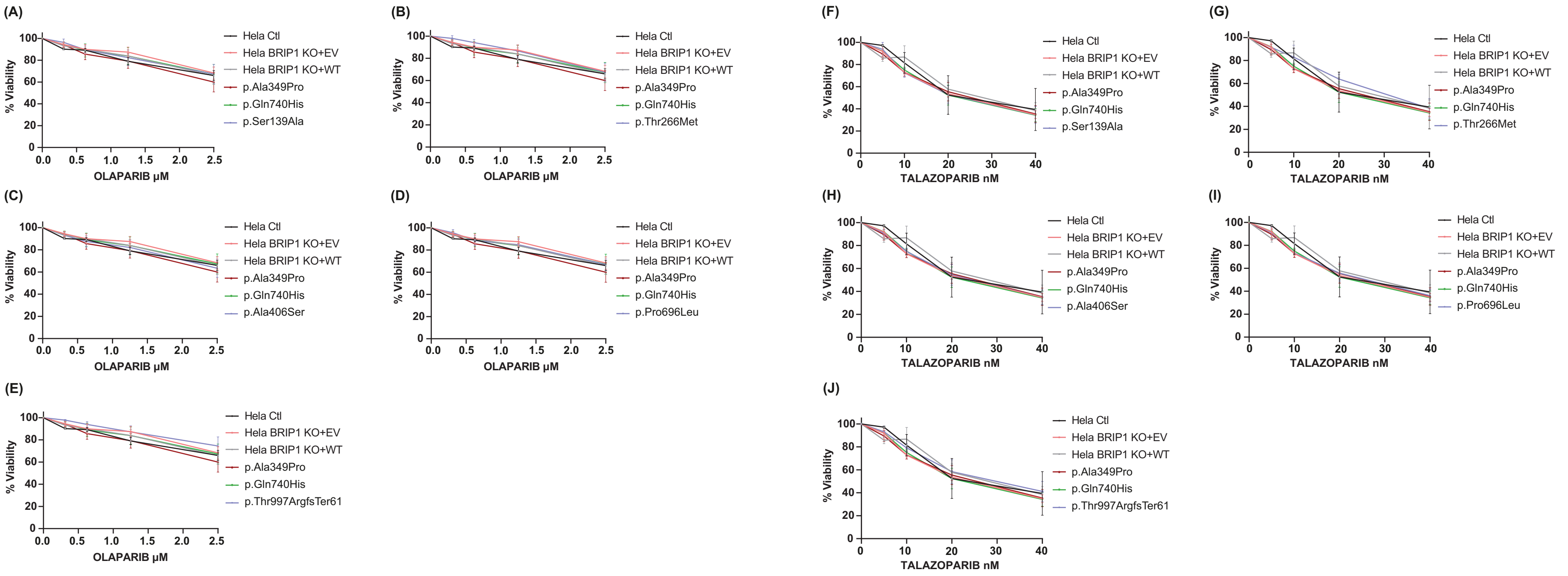


**Figure S4.** Sensitivity curves of BRIP1 variants to poly (ADP-ribose) polymerase inhibitors in Hela cells.
Survival curves contrasting the abilities of BRIP1 wild-type (WT) and the indicated variants, including the empty vector (EV), to rescue PARP inhibitors: olaparib (A-E) and talazoparib (F-J) resistance in Hela BRIP1-depleted cells.
